# A new content-qualified antenatal care coverage indicator: development and validation of a score using national health surveys in low- and middle-income countries

**DOI:** 10.1101/2020.02.28.20028720

**Authors:** Luisa Arroyave, Ghada E Saad, Cesar G Victora, Aluisio J D Barros

## Abstract

**Background:** Good quality antenatal care (ANC) helps reducing adverse maternal and newborn outcomes, especially in low and middle-income countries (LMICs). Most of the currently used ANC indicators only measure contact with services. We aimed to create and validate a new indicator measured as a score, considering both contact and content, that can be used for monitoring.

**Methods:** We used data from national surveys conducted in LMICs. Information on ANC was used to build an adequacy score (ANCq) that would be applicable to all women in need of ANC. Cronbach’s alpha and factor analysis were used to assess the proposed indicator. We also used a convergent validation approach, exploring the association of our proposed indicator with neonatal mortality.

**Findings:** The ANCq score was derived from seven variables related to contact with services and content of care ranging from zero to ten. Surveys from 63 countries with all variables were used. The validity assessment showed satisfactory results based on Cronbach’s alpha (0 82) and factor analysis. The overall mean of ANCq was 6 7, ranging from 3 5 in Afghanistan to 9·3 in Cuba and the Dominican Republic. In most countries, the ANCq was inversely associated with neonatal mortality and the pooled for all surveys OR was 0 90 (95%CI 0-88-0-92).

**Interpretation:** ANCq allows the assessment of ANC in LMICs considering contact with services and content of care. It also presented good validity properties, being a useful tool for assessing ANC coverage and adequacy of care in monitoring and accountability exercises.

**Funding:** Bill & Melinda Gates Foundation (Countdown to 2030), the Wellcome, Associação Brasileira de Saúde Coletiva (ABRASCO) and Coordenação de Aperfeiçoamento de Pessoal de Nível Superior (CAPES).

**Research in context:** *Evidence before this study:* Antenatal care (ANC) is an important part of primary health care, being associated with reductions in maternal and new-born morbidity and mortality, mainly in low and middle-income countries (LMICs). Several indicators have been proposed to measure ANC quality either through contacts with services or based on content of care, or sometimes both. We have reviewed the literature in PubMed, Web of Science and LILACS databases, using the terms “prenatal care”, “prenatal”, “antenatal care”, “care, antenatal”, “antenatal”, “atencion prenatal”, “cuidado pre-natal”, “Demographic and Health Surveys”, “DHS”, “Multiple Indicators Cluster Survey”, “MICS”, without any other restriction. Most of the proposed indicators that we found classified ANC quality in two or a few categories and are applicable only to women who had at least one ANC visit. Thus, women who did not receive any care and yet have a need for ANC are left out of those indicators. Even though there is consensus that an ANC indicator that includes aspects of quality of care is needed for monitoring ANC progress, none of the proposed measures has been widely adopted.

*Added value of this study:* We proposed a content-qualified ANC coverage indicator in the form of a score, called ANCq, which is applicable to all pregnant women and easily estimated using data from national surveys conducted in LMICs. It includes seven different variables related to contact with services and content of care received during pregnancy. The ANCq has good validity properties and was inversely associated with neonatal mortality. The novel indicator showed that there is wide variation across countries regarding ANC and also large within-country gaps.

*Implications of all the available evidence:* The proposed indicator provides a standardized and comparable measure of ANC adequacy, allowing for comparisons between and within countries. This indicator can help monitoring ANC progress to all women in need of ANC, with several advantages over currently existing indicators: it is applicable to all pregnant women independent of having accessed ANC services, it includes several aspects of ANC content and, being a score, provides a gradation of how suitable ANC was.

## Introduction

Antenatal care (ANC) is considered an essential part of primary healthcare during pregnancy, offering services that can prevent, detect and treat adverse maternal and newborn outcomes.^1–3^ Despite multiple efforts towards increasing coverage of ANC services and improve their quality, success has been limited in low and middle-income countries (LMICs), where maternal and neonatal mortality remain high.^3–5^ Further efforts are still required to achieve the 2030 agenda for Sustainable Development Goals (SDG), specifically target 3 that seeks to ensure healthy lives and promote well-being for all at all ages.

In 2016, the World Health Organization (WHO) updated the ANC guidelines, aimed at reducing the risk of stillbirths and pregnancy complications, and improving the ANC quality. The recommended number of ANC contacts was increased from four to eight, based on recent evidence indicating that a higher frequency of ANC contacts with a health provider is associated with a reduced likelihood of stillbirths.^4^ Most of the currently used ANC indicators for monitoring ANC in the context of the SDGs are based on a visit count. However, there is consensus in the literature that ANC quality should not be solely measured through the number of visits, but also include information on content of the care received, particularly regarding an essential set of interventions and assessments that are required for every pregnancy.^4,6^

Several authors have proposed different types of quality indicators for ANC.^1–3,5,7–11^ Some have proposed binary indicators^1–3^ or categorical classifications,^11,12^ considering the number of interventions received by pregnant women. In most studies, good ANC quality was defined as having received all or most of the components considered.^1,2,8,9^ A “quality index” was proposed by Dettrick et al ^10^ using principal components analysis to derive weights and calculate a score. Most of these indicators of ANC quality are restricted to pregnant women who had at least one ANC visit, thus leaving out those who did not receive any care, and yet have a need for ANC.^1,5^

Although there is consensus among researchers on the need for a comprehensive ANC quality indicator for monitoring progress, none of the proposed measures has been widely adopted. In this article we propose an ANC indicator in the form of a score that includes both contact with services and the content of care received during pregnancy. The indicator is applicable to all women in need of ANC and is estimated using data from national health surveys, the main source of health information for LMICs.

## Methods

We used data from Demographic and Health Surveys (DHS) and Multiple Indicator Cluster Surveys (MICS), which are nationally representative household surveys providing data on a wide range of health indicators with a focus in reproductive, maternal and child health. DHS and MICS use standardized data collection procedures across countries, comparable across and between surveys.^13,14^

We analyzed the most recent survey for each country with publicly available datasets, carried out since 2010. Data on ANC refers to the last child born to each woman aged 15-49 years. The recall period includes five years before the survey for DHS, and two years for MICS.

The rationale that guided us in building this new ANC indicator was:

- To create a single indicator including information on contact with health services and content of care received;
- To cover all women in need of ANC - as expected from the denominator of a coverage indicator – rather than restricting it to women with at least one ANC visit;
- Instead of a categorical indicator (e.g. “adequate” or “inadequate”), to develop a numerical score providing a measure of adequacy. A score ranging from zero to ten seemed the most intuitive;
- To group of the number of ANC visits into categories, based on current and previous WHO recommendations;
- To assign equal weights to all interventions, given that their importance may vary depending on the context, and also from woman to woman;
- To include component items that are deemed desirable in a good quality ANC, namely a first visit during the first trimester of gestation; at least one visit with a skilled provider and as many ANC-related interventions as possible in a way to maximize the number of surveys for which the indicator is applicable.

Our first step was to identify all questions related to ANC available in DHS and MICS, especially those related to content of care received, which are the most variable in the surveys (Table S1). Next, we determined the number of surveys with available information for each question in order to select those that could be used in the score (Table S2).

Using variables that are available in a large proportion of surveys, we gave arbitrary values to each ANC component, as described in Table 1.

**Table 1.**
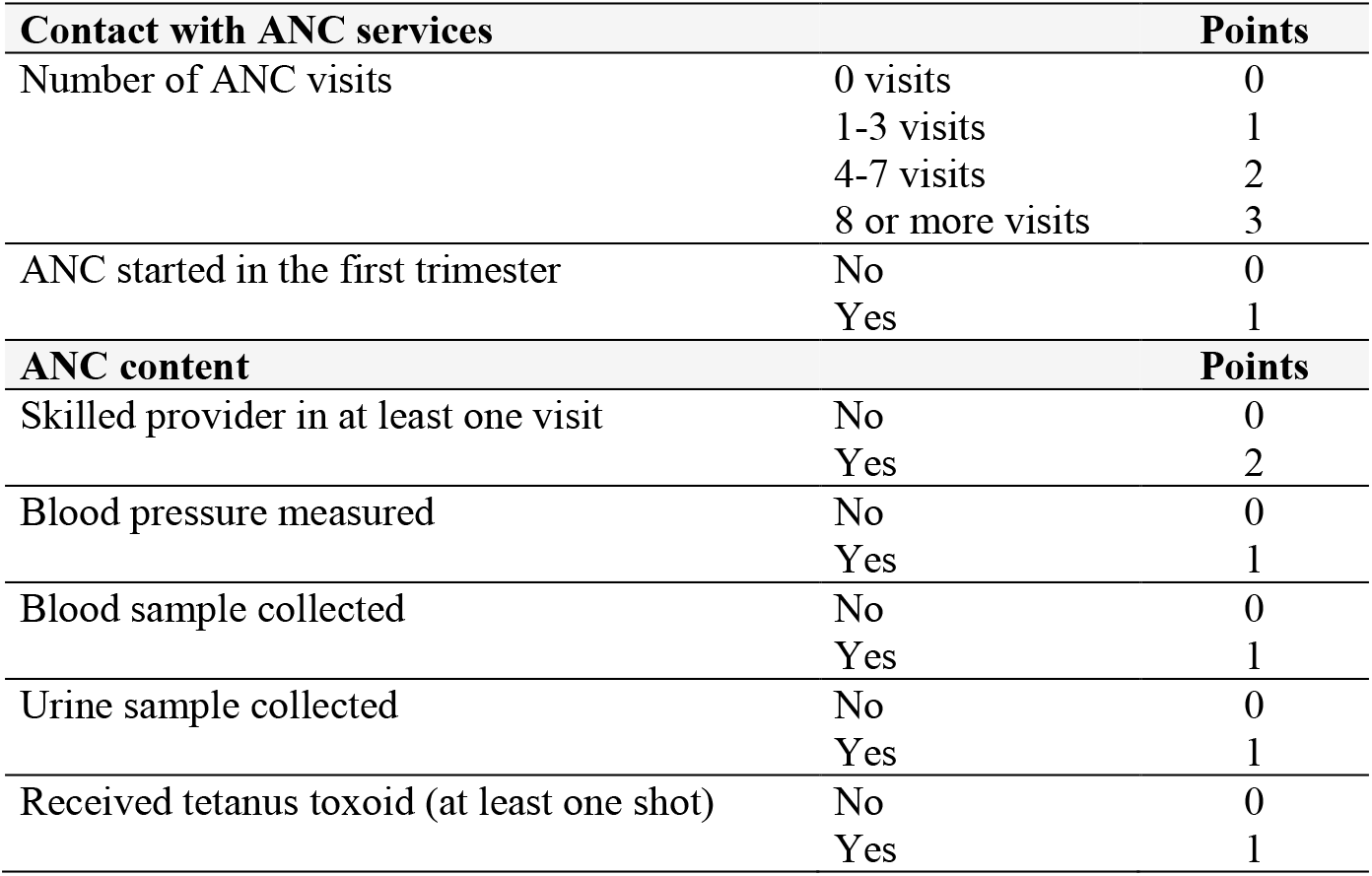
Scoring of the variables that compose the content-qualified ANC indicator, ANCq.

| **Contact with ANC services** |  | **Points** |
| --- | --- | --- |
| Number of ANC visits | 0 visits | 0 |
|  | 1-3 visits | 1 |
|  | 4-7 visits | 2 |
|  | 8 or more visits | 3 |
| ANC started in the first trimester | No | 0 |
|  | Yes | 1 |
| **ANC content** |  | **Points** |
| Skilled provider in at least one visit | No | 0 |
|  | Yes | 2 |
| Blood pressure measured | No | 0 |
|  | Yes | 1 |
| Blood sample collected | No | 0 |
|  | Yes | 1 |
| Urine sample collected | No | 0 |
|  | Yes | 1 |
| Received tetanus toxoid (at least one shot) | No | 0 |
|  | Yes | 1 |

We calculated Cronbach’s alpha to verify the internal consistency of our indicator. We also conducted confirmatory factor analysis^15^ to assess whether the indicator was compatible with a one factor solution and its goodness of fit. Given the non-normal nature of the variables, factor analysis was adjusted using robust maximum likelihood estimation. The standardized root mean squared residual and the coefficient of determination were evaluated. Standardized root mean squared residual measures the difference between the residuals of the sample covariance matrix and the hypothesized model while the coefficient of determination indicates how well the model fits.

In the absence of a gold standard to which our indicator could be compared, we carried out convergent validation exercise for external validity. It is widely accepted that a good quality ANC will reduce the risk of neonatal mortality.^11,16,17^ We therefore explored the association between our proposed indicator with this outcome, but we do not intend it to be a predictor of neonatal mortality.

Using the birth history recorded in the surveys, we defined neonatal death as that occurring during the first 30 days of life (the usual definition used in surveys given deaths occurring around the end of the first month are often reported at one month of age). For the neonatal mortality analysis, we only used DHS because these surveys allow linking birth history with ANC data. We included surveys with ten or more neonatal deaths and analyzed the last child born alive for the women in the previous five years.

We used logistic regression to analyze the relationship between our proposed score and neonatal mortality, estimating an odds ratio (OR) for each country. We then pooled all surveys using a random effects meta-analytic approach to obtain a pooled OR.

We also adjusted the models by household wealth, women’s age, and education in order to examine whether its effect was independent of these distal sociodemographic determinants. To allow for non-linearity in the association, we used a fractional polynomial approach to find the best fitting model for the pooled data.

Given that variables available for a small number of surveys were not used in our indicator, we conducted a sensitivity analysis to explore whether adding them would make any relevant difference in relation to our proposed score. We used principal component analysis (PCA) to create comparable scores for the various scenarios. We, then, calculated the correlation coefficients between the scores for the extended indicators and the score for our proposed set of variables. We also estimated the association between each resulting score and neonatal mortality to assess differences.

Finally, we compared the performance of our indicator in predicting neonatal mortality with other existing indicators in the literature that were applied for a set of surveys and not just for a specific country (Table S4). This was done to check whether any of the indicators has a clear predictive advantage. For that, we calculated the area under the ROC curve (AUC) for each indicator along with its confidence interval.

The analyses were performed using Stata 16.0 (StataCorp, College Station, TX), always taking into account the survey design (clustering and sampling weights).

The study was based on anonymized publicly available data. Therefore, the analyses did not require ethical clearance. This was done by each of the institutions responsible for carrying out the original surveys. Patients or the public were not involved in the design, conduct, reporting, or dissemination plans of our research.

## Results

We identified 99 nationally representative surveys carried out since 2010. Seven variables related to ANC coverage and quality were present in 63 surveys and were selected to compose the indicator. The full set of variables considered is presented in Tables S1 and S2 (supplementary material). Two of the seven variables were related to contact with services (timing of the first visit and the total number of visits) and five were related to content of care (at least one visit with a skilled provider, blood pressure measurement, blood and urine samples collection, and administration of at least two shots of tetanus toxoid).

The 63 surveys (42 DHS and 21 MICS) were conducted between 2010-2017 in LMICs from six UNICEF world regions. In total, we studied 583,602 women with a live birth in the five (DHS) or two (MICS) years before the survey.

The proposed score, named ANCq, ranges from zero to ten points. Table 1 shows that each variable was coded as zero or one, except for number of visits (categorized according to previous and current WHO recommendations) and being seen by a skilled provider (zero for “no” and two for “yes”), given the relevance of provider for ANC quality. Providers considered as skilled included doctors, midwives, nurses, and other attendants considered as skilled by each country, such as auxiliary midwives.

A pooled dataset with the 63 surveys with suitable data was put together for the analyses. The validity assessment of the indicator showed satisfactory results, with Cronbach’s alpha coefficient equal to 0·82. The confirmatory factor analysis indicated that a single factor solution was adequate, with the first factor presenting an eigenvalue of 3·68 and explaining 5·25% of the total variance. All other factors had eigenvalues below one, the usual cut off value for selecting relevant factors. The loadings of the variables ranged from 0·31 for tetanus injection to 0·84 for blood pressure measure, all above the recommended minimum of 0·30 for loadings. The confirmatory analysis indicated the model fitted the data well with a standardized root mean squared residual = 0·05 (values less than 0·08 are recommended) and a coefficient of determination = 0·886 (maximum value of one).

In order to give an idea of the percentage of women in each category of the variables that are part of the indicator, we present the median values (and interquartile ranges) across all 63 countries in Table 2. The median percentage of women attending four to seven ANC visits was 49·8%. The median percentage of women receiving care from a skilled provider for at least one visit was 95.8% and the lowest value observed was 54·9% for having started ANC in the first trimester.

**Table 2.** Median and interquartile range for the country estimates using DHS and MICS surveys from 63 low- and middle-income countries. Source: DHS and MICS, 2010-2017.

| Variable | Median | IQR |
| --- | --- | --- |
| **Number of ANC visits** |  |  |
| Zero visits | 2.5 | 0.1 - 6.5 |
| 1-3 visits | 22.9 | 9.2 - 36.3 |
| 4-7 visits | 49.8 | 32.8 - 57.7 |
| 8 or more visits | 13.7 | 3.9 - 35.2 |
| **ANC with skilled attendant** |  |  |
| Yes | 95.8 | 89.4 - 98.9 |
| **ANC started in first trimester of pregnancy** |  |  |
| Yes | 54.9 | 37.6 - 70.6 |
| **Blood pressure measured in ANC visit** |  |  |
| Yes | 92.5 | 79.3 - 96.8 |
| **Blood sample taken in ANC visit** |  |  |
| Yes | 85.9 | 67.0 - 94.5 |
| **Urine sample taken in ANC visit** |  |  |
| Yes | 82.5 | 53.7 - 93.4 |
| **2+ tetanus injections before birth** |  |  |
| Yes | 55.56 | 40.1 - 62.8 |

The distribution of the ANCq scores for all countries pooled together is presented in Figure 1. The overall mean score was 6·7. For 54·9% of the women, the score ranged from seven to nine points, with 8 and 9 being the most frequent values (approximately 20% each). The overall proportion of women with no ANC was 6·9%. Figure 1 also shows the distribution of each contact and content variable, according to the ANCq score in points. Women with one point in the score received mainly tetanus toxoid (97%), even though they did not attend ANC.

**Figure 1.**
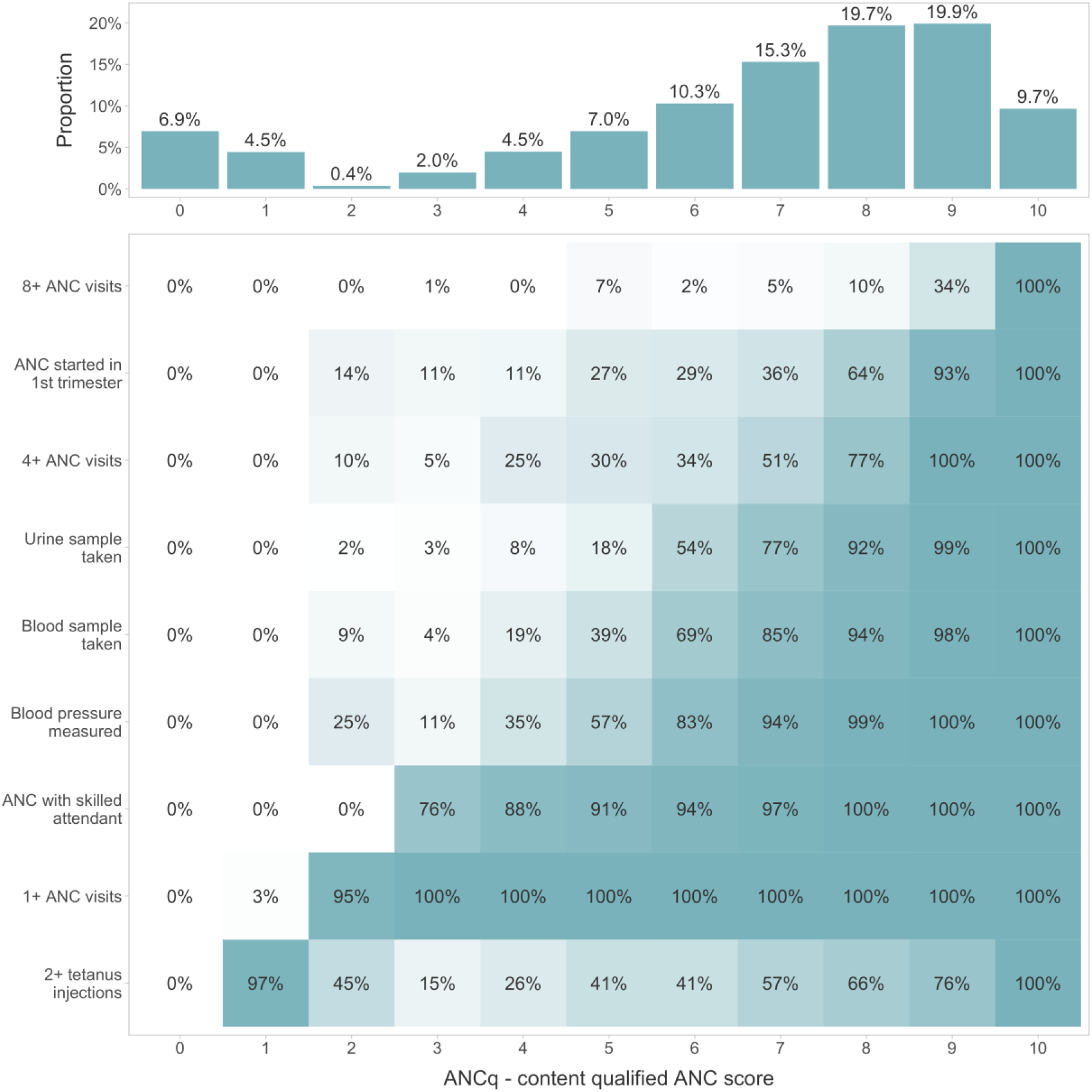
ANCq score distribution using DHS and MICS surveys from 63 low- and middle-income countries. Source: DHS and MICS, 2010-2017.

The country specific means of ANCq ranged between 3·5 for Afghanistan to 9·3 in Cuba and the Dominican Republic. Figure 2 presents box and whisker plots for ANCq by country, grouped by UNICEF world region. There is wide variation in ANCq within countries, between countries and between regions. Table S3 in the supplementary material presents the means and quartile cut-off points for each country.

**Figure 2.**
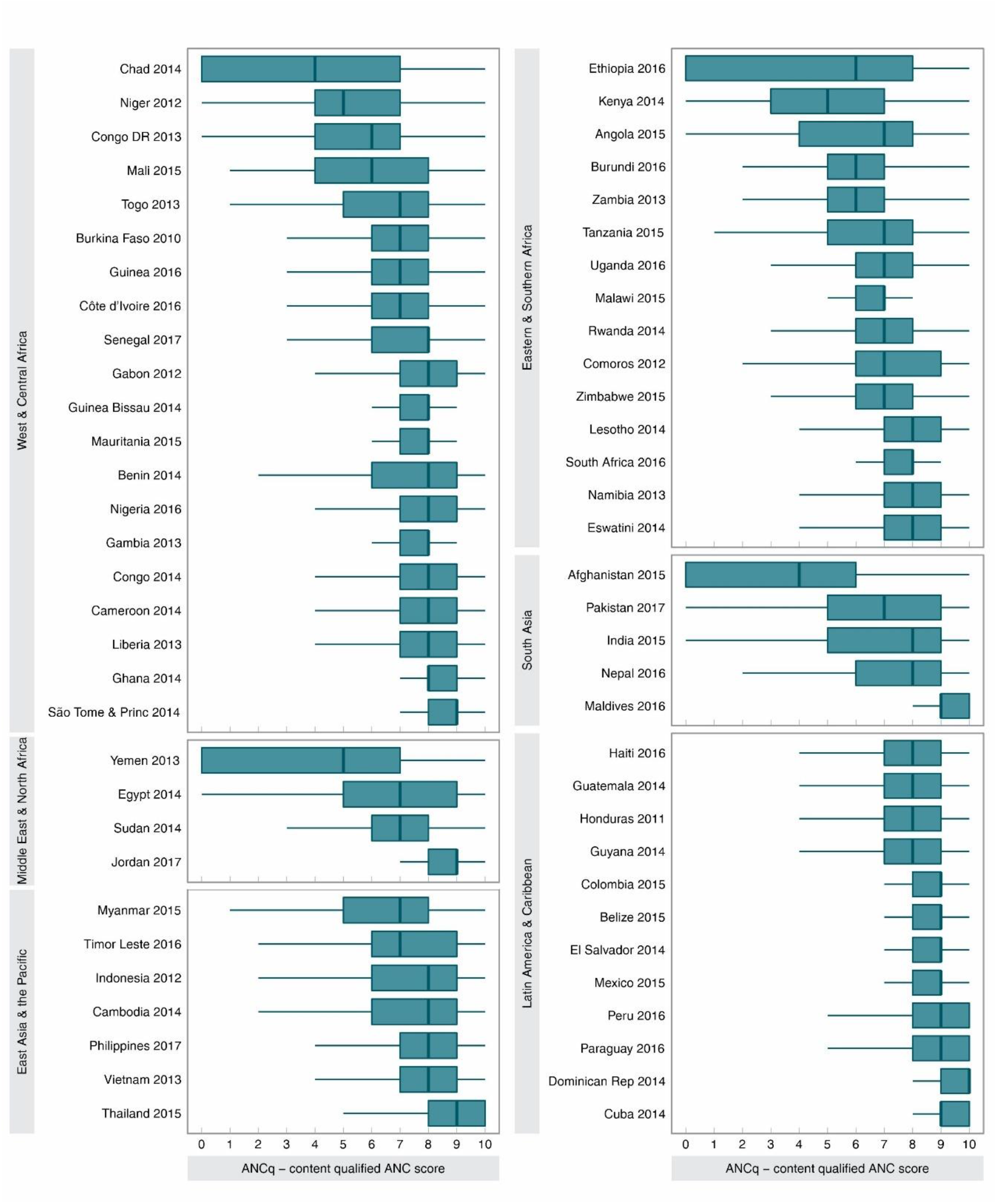
ANCq score distribution for each country grouped by UNICEF regions of the world. Source: DHS and MICS, 2010-2017

To explore how the ANCq score associated with neonatal mortality in each country, we fitted a simple logistic model separately for the 42 DHS surveys. In 27 countries the ORs were consistent with protection, given that their confidence intervals did not include the unity (Figure S1). In 11 countries, the ORs were below one, and in four countries above one, but in all these cases the confidence intervals included the unity. The pooled OR showed that each additional point in the score reduces the odds of neonatal mortality by 10% (OR: 0.90; 95% CI: 0·88-0·92). There was moderate heterogeneity between countries (I^2^: 60 2%). Adjusting the model for wealth, maternal age and education had a very small impact on the estimated OR, and it remained inversely associated with neonatal mortality (pooled adjusted OR: 0·92; 95% CI: 0·91-0·93).

Figure 3 shows the shape of the association between ANCq and the outcome using the pooled dataset through logistic regression and a fractional polynomial approach. We observed that the drop in mortality rate from score zero to one was the largest, followed by progressive declines following closely a straight line. On average, the neonatal mortality rate predicted for women with zero score was 33 deaths per thousand live births, whereas a rate of ten per thousand was predicted for those with the maximum score of ten.

**Figure 3.**
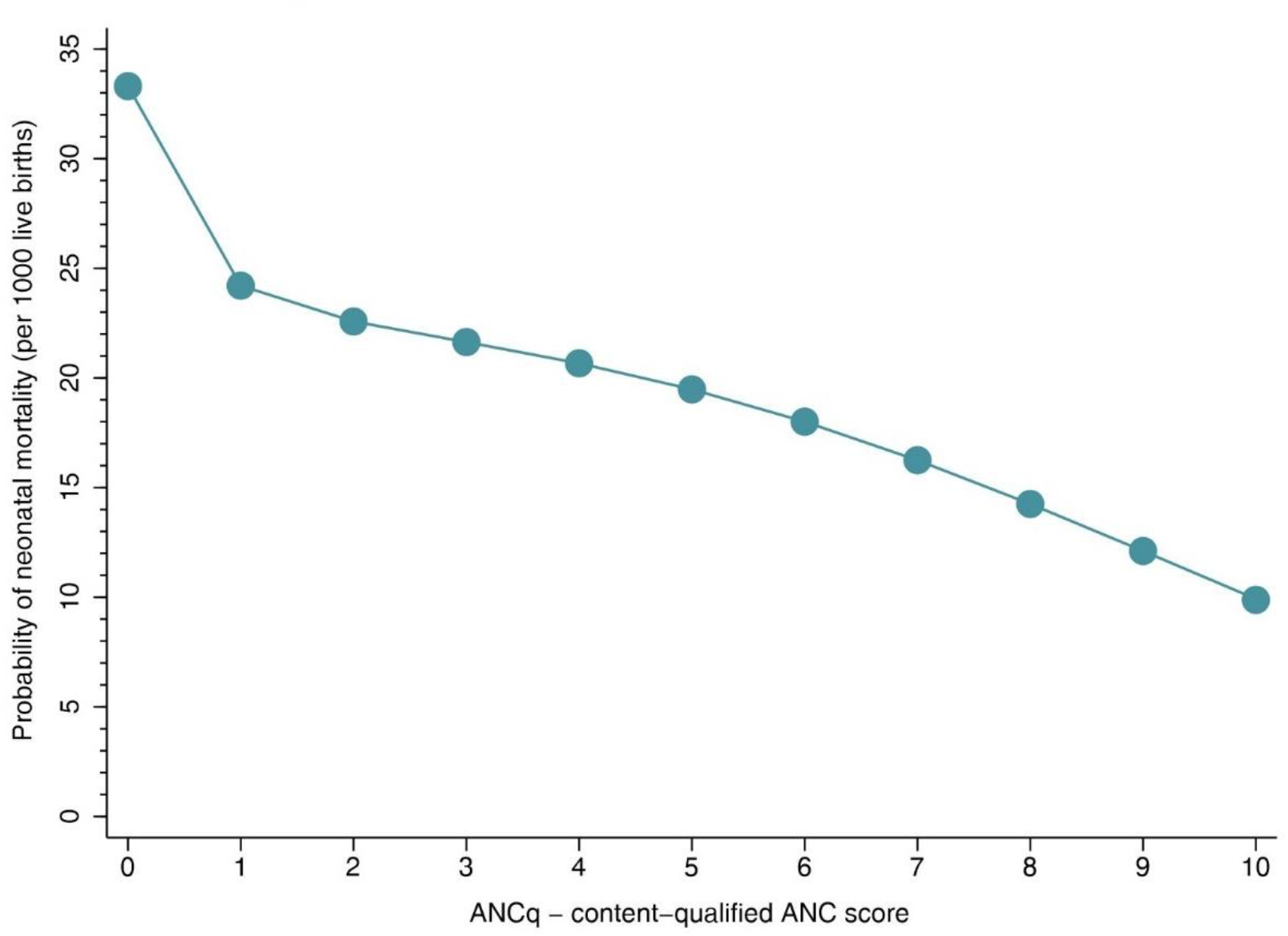
Predicted probabilities of dying in the first 30 days of life (neonatal mortality, as usually reported in surveys) according to the ANCq score. Source: DHS, 2010-2017.

In the sensitivity analysis, we compared five different sets of variables, each representing a potential alternative to our proposed ANCq. One set included the same variables in the ANCq. The other four sets added iron supplementation (90 days or more), information about pregnancy complications, weight and height measurement, HIV tested and HIV results. Each set with additional variables was estimable in a smaller number of surveys (Table S5). The correlation coefficients between the PCA scores obtained with the ANCq set of variables and the extended ones (Table S6) varied between 0·89 (with the largest set of variables, estimable in only four surveys) and 0·95. The ORs of the association of each score with neonatal mortality varied little, and all confidence intervals overlapped (Table S6).

Finally, we compared the ANCq and three other indicators identified from the literature in terms of their predictive power of neonatal mortality using the area under the ROC curve. ANCq presented the highest AUC (0·58; 95% CI: 0·57-0·59), followed by the indicators by Amouzou et al.^17^ (0·57; 95% CI: 0·56-0·57), Arsenault et al. ^5^ (0·52; 95% CI: 0·51-0·53) and Carvajal ^8^ (0·50; 95% CI: 0·50-0·50).

## Discussion

We proposed an ANC score indicator that comprises both contact with health services and content of care that was estimated for 63 countries, using national surveys. We also favored a more nuanced, numerical score rather than a categorical indicator, for which higher scores were associated with lower neonatal mortality, suggesting that the indicator is capturing relevant aspects of ANC. We believe that the development of a graded indicator which can be estimated from health surveys offers a powerful tool for monitoring progress in ANC, including aspects related to adequacy of care, in the context of the SDGs.

ANCq presented wide variation between and within countries. Latin America and the Caribbean was the region with higher average scores and less variability between countries. Although our results show that globally more than half of women scored between seven and nine points (55%), 7% received no care during pregnancy, which may be explained by contextual and individual factors.^18,19^ A systematic review showed that maternal education, household income, cultural belief and place of residence have an important influence on ANC coverage in LMICs.^20^

Our study has some limitations that should be noted. Whereas the surveys are nationally representative and comparable in terms of sampling strategy and data collection methods,^14^ there is ample variability in the information collected on ANC, and in most cases, the information on content is limited to a few variables. Specifically, many MICS lacked information on iron supplementation, one of the key ANC interventions. In order to estimate ANCq for a larger set of countries, iron supplementation information had to be excluded from ANCq, likewise, several other evidence-based ANC interventions were left out.^21^ As a result, the score may overestimate ANC adequacy, however, the sensitivity analysis results showed very high correlations between ANCq and expanded indicators, including a larger set of variables. This strongly suggests that adding more variables to ANCq will not imply any important change in how it assesses ANC. On the other hand, each additional variable, at least at this moment, makes ANCq not estimable for a large number of surveys, which is a serious problem in monitoring exercises.

Another limitation is that the information is based on self-report, and for DHS this may refer to care received during a pregnancy that took place up to five years before the survey. We are aware that data for children born in the two or five years preceding the survey can be affected by recall bias. Nevertheless, all survey-based pregnancy and delivery care indicators largely used for SDG monitoring suffer from the same problem. Finally, the change in WHO recommendation for the desired number of ANC visits, in 2016, is unlikely to have influenced the comparison of countries since only four surveys were done after that and they include births in the five years previous to the interview date.

The decision to attribute points to each item arbitrarily is debatable. Our starting point was to give equal weights to all available evidence-based interventions - since it is difficult to assess their relative importance - and to give higher weight to the number of visits and the type of provider. Most other studies measuring the ANC quality through scores also gave arbitrary weights for each item, and most often the same weight for each intervention included.^22,23^ Others relied upon data driven approaches such as PCA,^10^ but this ignores any theory in terms of the weights assigned. A notable result from our sensitivity analysis was the association between ANCq and neonatal mortality, which was stronger than the score obtained by PCA, where no arbitrary decisions are made in terms of weighting of variables. That would suggest that our theoretical understanding of the variables used works better than a purely data driven analysis. We started with a theoretical construct, and then showed that it was consistent with principal components results. The validity assessment of ANCq through Cronbach’s alpha and confirmatory factor analysis also presented satisfactory results.

The loading for tetanus injection before birth was considerably lower compared to the other variables. One possible reason is that it is possible to receive tetanus immunization outside the context of ANC visits, and also that its indication during pregnancy is also determined by past history of immunization. Unfortunately, this cannot be ascertained with the information available in the surveys. Despite the weaker loading for this variable, we decided to keep it in our indicator given its importance in preventing neonatal tetanus.

Most published studies on ANC quality were conducted using a single survey, which has the advantage of including a larger number of quality indicators according to national recommendations.^1,3,23,24^ However, this approach does not lend itself for a global monitoring indicator. The study by Arsenault et al. analysed 91 national surveys,^5^ but to do so the authors only included three ANC interventions – blood pressure checked and urine and blood sample collected – thus rendering the indicator less representative of what is perceived as adequate care. In other studies, the ANC quality was analyzed – either as an outcome or exposure variable – using selected surveys. However, only a few DHS and MICS surveys have information for all variables included in those proposed indicators (Table S4). Lastly, most studies have completely left out pregnant women who did not have any ANC visits, and therefore did not measure population coverage. Emulating these indicators for our surveys, we showed that ANCq has better or comparable predictive power for neonatal mortality than those proposed in multicountry analyses what reinforces its quality as a potential coverage indicator in monitoring exercises.

Studies of ANC quality among attenders^5,8^ are well suited to answer the question of quality of services and have a place in the quality literature. We explicitly chose to propose a coverage indicator with all women in need of ANC in the denominator, that would also include aspects of ANC content, and thus head in the direction of measuring effective coverage (EC). In general, EC indicators involve the quality of service and not just receiving it.^25^ However, many definitions and methods have been used, varying across the literature. Some authors have used health facility data or data from facility based surveys to measure the adequacy of care or the EC.^26–28^ These surveys collect information on staff, infrastructure, resources, procedures and support systems available, as well as the satisfaction for patient and service providers.^29,30^ This is a powerful approach but it does not allow for individual level assessment, which is essential when doing equity analysis, since these analysis preclude subgroup comparisons.

Given the lack of a gold standard indicator for ANC quality in surveys, we resorted to a convergent validation strategy. Neonatal mortality was chosen as the available outcome known to be related to ANC. We showed that ANCq was inversely and monotonically associated with it - the higher the score, the lower the associated risk. Similar associations have been reported elsewhere.^16,17^ One study from Zimbabwe reported reduction of 42·3%, 30·9% and 28·7% in neonatal, infant and under-five mortality, respectively, for children whose mothers received good ANC quality.^16^ Another study including data from 60 LMICs showed a 18% reduction in neonatal mortality among babies of women who had adequate ANC.^11^ It is noteworthy that we explored the association between ANCq and neonatal mortality in order to conduct a convergent validation strategy. We did not want to create a predictor of mortality with the ANCq.

In conclusion, we have proposed a valid indicator that covers several aspects of ANC, which is easily estimated using survey data from LMICs, and also allows for trend analyses. It goes beyond the traditional ANC indicators that mostly rely on contact with service and avoids the use of multiple indicators to assess ANC services in a given setting, being a useful addition to the arsenal of indicators of the health related SDGs.

## Data Availability

The original datasets from DHS and MICS are freely available

http://dhsprogram.com/

http://mics.unicef.org/

## Acknowledgements

We thank the Bill & Melinda Gates Foundation, the Wellcome Trust, Associação Brasileira de Saúde Coletiva and Coordenação de Aperfeiçoamento de Pessoal de Nível Superior (CAPES) for funding this study. We are thankful to Cintia Borges and Thiago Melo for your help in the graphic design.

## Contributors

LA and AJDB conceptualized the paper and conducted the analyses, with support from CVG and GES. LA interpreted the results and wrote the manuscript with technical support from AJDB. AJDB, GES and CGV contributed to critically review the analysis and writing. AJDB originally proposed the idea of the indicator and advised on the analysis. All authors read and approved the final manuscript.

## Patient consent for publication

Not required.

## Data availability statement

The original datasets from DHS (http://dhsprogram.com/) and MICS (http://mics.unicef.org/) are freely available.

## Open access

This is an open access article distributed in accordance with the Creative Commons Attribution Non Commercial (CC BY-NC 4.0) license, which permits others to distribute, remix, adapt, build upon this work non-commercially, and license their derivative works on different terms, provided the original work is properly cited, appropriate credit is given, any changes made indicated, and the use is non-commercial. See: http://creativecommons.org/licenses/by-nc/4.0/.

